# A prospective observational cohort study considering the feasibility and tolerability of high intensity interval training as a novel treatment therapy for patients with intermittent claudication

**DOI:** 10.1101/2020.04.01.20049569

**Authors:** Sean Pymer, Said Ibeggazene, Joanne Palmer, George E. Smith, Amy E. Harwood, Sean Carroll, Lee Ingle, Ian C. Chetter

**Affiliations:** Academic Vascular Surgical Unit Hull York Medical School, Hull, UK; Discipline of Sport and Exercise Science Faculty of Medicine and Health Sciences, University of Sydney, Sydney, Australia; Department of Sport, Health & Exercise Science, University of Hull, Hull, UK

**Keywords:** Intermittent Claudication, Supervised Exercise Programmes, High-Intensity Interval Training, Peripheral Arterial Disease, Vascular Surgery

## Abstract

**Background:** Uptake and completion rates for supervised exercise programmes (SEP) for patients with intermittent claudication (IC) are low. Current exercise prescription is a *one size fits all approach*, based on claudication pain thresholds, potentially limiting individual benefits. High-intensity interval training (HIIT) has the potential to overcome these barriers as it is a more time-efficient, objectively prescribed exercise programme. This study aimed to assess a novel HIIT programme for patients with IC in terms of safety, feasibility, tolerability and indicators of efficacy.

**Design/Methods:** Patients referred to a usual-care SEP were invited to undertake an alternative HIIT programme. All recruited patients performed a baseline cardiopulmonary exercise test (CPET), to inform their exercise prescription. HIIT involved ten, one-minute high-intensity intervals on a stationary cycle ergometer interspersed with one-minute recovery intervals, performed three times per week for six-weeks. Outcomes included safety, feasibility, tolerability, walking distance and quality of life (QoL).

**Results:** 144 patients with IC were referred, 95 met initial eligibility criteria (66%) and 30 (32%) were recruited for HIIT, of which 15 (50%) completed. Of the recruited patients, 90% were on optimal medical therapy and 40% had concomitant cardiac, cerebrovascular and/or respiratory disease.

One serious adverse event was recorded and patients who completed the programme attended 100% of the sessions. Improvements in walking distances and quality of life were observed. Following recruitment of the first 20 patients, the inclusion criteria was refined on the basis of CPET, leading to improved completion rates.

**Conclusion:** The study provides preliminary findings indicating that patients with IC can complete a short-term HIIT programme. HIIT also appears safe, well-tolerated and although not formally powered, walking distances and QoL appear to improve following HIIT. Further research to evaluate the role of HIIT in patients with IC seems warranted.

## Introduction

Peripheral arterial disease (PAD) is characterised by atherosclerosis of the arteries supplying the lower limbs, resulting in a reduced blood supply ^1^. PAD is an increasingly prevalent disease, estimated to affect 202 million people globally, with an increase of 23.5% between the years 2000 and 2010 ^2^. The most common symptom of PAD is intermittent claudication (IC) ^3,4^. IC is characterised by reproducible ischaemic muscle pain in the leg, precipitated by exertion and relieved by rest, due to an oxygen demand-supply imbalance ^3,5-7^. IC has a deleterious effect on walking ability, functional capacity, quality of life (QoL), daily activities and balance, whilst also carrying a markedly increased premature mortality risk ^3,5,8-13^. First line management of IC, recommended by national and international guidelines, includes pharmacological cardiovascular risk factor optimisation and exercise therapy ^4,14^. Exercise therapy should ideally involve a supervised exercise programme (SEP), consisting of two hours of exercise per week, for a 12-week period whereby patients are encouraged to exercise to the point of maximal lower leg pain ^14^. SEPs are more beneficial than unsupervised programmes ^15,16^, with the level of supervision being related to the improvement in walking distance ^17^. Interval walking is the recommended exercise component of SEPs, though improvements are also obtainable with a variety of other modalities, including stationary cycling ^18,19^. In addition, cycle testing is better tolerated than treadmill testing in IC patients, whilst also inducing greater cardiometabolic responses ^20^, likely because patients may prematurely terminate treadmill testing due to claudication pain, precluding them from reaching higher intensities ^20^.

The evidence for the efficacy of SEP, irrespective of exercise training modality, is irrefutable, with a recent Cochrane review demonstrating it provides significant improvements in both intermittent claudication distance (ICD) and maximum walking distance (MWD) ^18^. Despite the overwhelming evidence of clinical and symptomatic benefits of SEP, less than half of vascular units in the UK have access to such programmes ^21^. Additionally, SEPs seem unpopular with patients, with recruitment rates often as low as 25% ^22^. Patients often cite time and SEP duration as reasons for inability to attend ^23^.

Despite its proven effectiveness, the exercise prescription within a SEP usually adopts ‘*a one size fits all’* approach based on a subjective measure of claudication pain. This may potentially limit the benefits that could be attained. High-intensity interval training (HIIT) is a personally prescribed exercise intervention based on objectively measured cardiorespiratory fitness via cardiopulmonary exercise testing (CPET). It therefore has the potential to maximise individual benefit. Additionally, HIIT is more time efficient, providing shorter individual exercise sessions within a programme, which may also overcome patient cited time barriers. HIIT has demonstrated similar or superior physiological benefits than lower intensity programmes across both healthy and clinical populations ^24-29^. HIIT has also been shown to elicit greater enjoyment than lower intensity alternatives and may be a preferred treatment option for IC patients ^23,30^. A recent systematic review has provided initial, limited evidence that HIIT may be beneficial for the IC population whilst recommending further studies of low-volume, short-term HIIT ^31^. Therefore, the aim of this study was to assess a novel HIIT programme for patients with IC in terms of safety, feasibility, tolerability and indicators of efficacy.

## Methods

### Study design

This was a prospective observational cohort study conducted at a tertiary vascular centre in the United Kingdom. Approval was obtained via a local NHS research ethics committee (Bradford Leeds – 18/YH/0112) and all patients provided written informed consent prior to participation. The study was registered on clinicaltrials.gov (NCT04042311).

### Participants and Recruitment

This was a prospective observational cohort study conducted at a tertiary vascular centre in the United Kingdom. Approval was obtained via a local NHS research ethics committee (Bradford Leeds – 18/YH/0112) and all patients provided written informed consent prior to participation. The study was registered on clinicaltrials.gov (NCT04042311).

### Participants and Recruitment

Patients diagnosed with IC by a team of consultant vascular surgeons and referred for our conventional SEP, were screened for study participation. Patients were deemed eligible if they were aged >18 years, English speaking and able to follow exercise instructions, able to provide informed consent, walk unaided and had a resting ABPI < 0.90 or a reduction of ≥ 20 mmHg following exercise testing. Those who had critical limb threatening ischaemia (rest pain and/or tissue loss), were undergoing active cancer treatment, had inadequately controlled cardiometabolic diseases, or elicited any contraindications to exercise testing or training were excluded ^32^. Patients meeting these criteria who were willing to participate were invited to attend a baseline assessment which included a CPET. Following this test, patients were also excluded based on significant exercise-induced myocardial ischaemia, manifesting as significant ECG changes and/or an abnormal haemodynamic response to maximal exertion. In addition, as achieving a maximal effort CPET is required for accurate and effective *traditional* HIIT exercise prescription, patients were initially excluded if they were unable to achieve maximal effort criteria ^33,34^, including combined self-perceived exertion and physiological measures (see table 1) ^33,34^. However, following a review of the inclusion/exclusion criteria due to a completion rate that was lower than anticipated for the first 20 patients, this maximal effort criterion was removed and no longer applied to the final 10 patients that make up the 30 patients in this cohort.

**Table 1.** Adopted maximal effort criteria ^33,34^

|  |
| --- |
| A Plateau in $\dot{V}O_2$ (or failure to increase by 150 ml/min) with an increased workload. (NB. This is rarely observed in a patient population) |
| Achievement of >85% of age-predicted maximal heart rate |
| A respiratory exchange ratio ($\dot{V}O_2/\dot{V}CO_2$) at peak exercise $\geq 1.10$ |
| An RPE of >17 on the 6-20 Borg scale |
| $\dot{V}O_2$ = oxygen uptake, $\dot{V}CO_2$ = carbon dioxide production, RPE = rating of perceived exertion.<br>N.B. Attainment of two or more of these criteria was required for achievement of maximal effort to be considered. |

### Outcome measures

The primary aims of this study were to assess a short-term, low-volume HIIT intervention in patients with IC in terms of safety, feasibility, tolerability, and indicators of efficacy.

Safety assessment included; adverse and serious adverse event reporting related to the intervention/outcome measures.

Feasibility assessment included; eligibility rates (number of patients eligible / screened), recruitment rates (number of patients recruited / eligible) and completion rates (number of patients completing the HIIT programme / recruited).

Tolerability assessment included; assessment of reasons for withdrawal (i.e. whether or not they were related to the exercise training intervention) and the ability of patients to achieve and maintain exercise in the appropriate exercise intensity domain (i.e. ≥85 % peak heart rate), for 10 full HIIT intervals.

Indicators of clinical efficacy assessment included; graded walking treadmill test (to assess ICD and MWD), pre and post exercise ABPI, generic and disease specific QoL and cardiorespiratory fitness measurements. The indicators of clinical efficacy were assessed at baseline and immediately following completion of the HIIT programme. These measures were analysed descriptively rather than statistically, as this study was not appropriately powered to detect a statistically significant difference.

### Procedures

Ankle brachial pressure index (ABPI) was recorded at rest using a handheld Doppler (SummitDoppler, Wallach surgical devices, Trumbull, USA) and sphygmomanometer. Systolic blood pressure was measured in the left and right brachial, dorsalis pedis and posterior tibial arteries and the ABPI was calculated by dividing the highest of the two ankle pressures on each side with the highest of the two arm pressures. After resting ABPI was recorded, patients underwent a graded treadmill walking test, with a constant speed of 3.2km/h and incremental gradient starting at 0% and increasing by 2% every two minutes, for a maximum of 15 minutes ^35^. Those unable to walk at 3.2km/h were permitted to walk at a slower speed, selected by the assessor, which was kept consistent at follow up to ensure standardisation. ICD and MWD was recorded as the distance which patients first reported claudication pain and the distance at which pain became too severe and they needed to stop. Post-exercise ABPI was then recorded using the same methodology outlined above.

### Quality of Life

Both generic and disease specific QoL were measured using the Medical Outcomes Study Short Form 36 version 2 (SF-36) and the King’s College Hospitals Vascular QoL (VascuQoL) questionnaires ^36,37^. Both questionnaires have several domains with the SF-36 including physical functioning, social functioning, bodily pain, mental health, vitality, role physical, role emotional and general health and the VascuQoL including pain, activities, symptoms, emotion and social activities. Physical and mental component QoL scores can be derived from the SF-36, whilst a total score can be derived from the VascuQoL ^36,37^.

### Cardiorespiratory fitness

All patients performed a symptom-limited ramp incremental cycle ergometer (Lode Corival Serial, Groningen, Netherlands) CPET. Breath-by-breath metabolic gas exchange data were collected using a metabolic cart (Ultima2, Medgraphics, St Paul, Minnesota, USA), which was calibrated to manufacturers’ instructions prior to each test. Patient’s heart rate (HR) and rhythm was monitored continuously via 12-lead ECG (Mortara X-scribe, Mortara, Milwaukee, USA). Oxygen saturation (SpO_2_) and brachial blood pressure (BP; Tango M2 system, SunTech Medical, Morrisville, USA) were assessed periodically throughout the exercise test.

Each CPET was preceded by a 3-minute rest period, whereby patients were seated on the bike, to collect resting ECG, HR and BP data. This was followed by a 3-minute unloaded phase which preceded a set patient-specific ramp protocol of 10, 15, or 20 Watts per minute, designed to induce volitional exhaustion within 8-12 minutes ^38^. Patients were instructed to maintain 60-70 revolutions per minute (rpm) and in the absence of clinical indications for stopping, participants were encouraged to exercise until volitional exhaustion. The Borg 6 to 20 rating of perceived exertion (RPE) scale was used to quantify degree of effort (38). The requirement to give a maximal effort was explained thoroughly and strong verbal patient encouragement was given throughout. HR, RPE and SpO_2_ were recorded every minute and BP recorded every two minutes. Expired ventilatory gases were continuously collected during the rest and exercise periods and for at least 6 minutes into recovery ^39^. Tests were terminated if the patient exhibited any termination criteria as previously outlined ^32^.

Data were exported using 5s, 30s and 8 breath mean averaging for offline analysis. Peak oxygen uptake 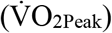 was defined as the highest value achieved during the last 30s of exercise or early in recovery. The ventilatory anaerobic threshold (VAT) was determined using the V-slope method, verified with the ventilatory equivalents ^40,41^.

### Intervention

The HIIT intervention was designed based on a large ongoing randomised controlled trial in patients with coronary artery disease ^34^, and consisted of 6 weeks of supervised, intermittent cycling 3 days per week, prescribed on the basis of the baseline CPET. Patients completed 10 × 1-minute high-intensity intervals (set at 85-90% peak power output [PPO] from CPET, to achieve ≥85% peak heart rate [HRpeak]) interspaced with 1-minute recovery intervals (20-25% PPO) on a Wattbike Trainer (Wattbike, Nottingham, UK), to achieve a total of 20 minutes exercise time. For pragmatic reasons, patients were permitted to complete less than 10 intervals, progressing up to 10 bouts on an individual basis. The variation in intensity was achieved by altering cycling cadence and patients were continuously monitored via a Polar heart rate monitor (FT2, Polar electro, Kempele, Finland) and RPE ^42^, with both recorded at the end of each high-intensity interval. The session was preceded and followed by a 10-minute warm-up and cool-down as is recommended for patients with cardiovascular disease ^43^.

### Data analysis

The primary aims of this study were to consider the safety, tolerability and feasibility of a novel HIIT programme. Therefore, descriptive rather than statistical analysis was performed. Data is presented as mean/mean difference ± standard deviation. In addition, due to the nature of this study and analysis, no formal sample size calculation is required.

## Results

### Patient Characteristics

Baseline characteristics of all recruited patients (*n* = 30) are shown in table 2. Mean age was 69 ± 9 years, body mass index was 29 ± 4 kg/m^2^, and 77% were male. The mean ABPI was 0.75 ± 0.21. Participants were typically (90%) on optimal medical therapy (i.e. statin and antiplatelet therapy) and only 30% required beta-blocker treatment. Concomitant cardiac, cerebrovascular and/or respiratory disease comorbidities were evident in 40% of participants at baseline. Exercise programme completers and non-completers were similar in baseline characteristics.

**Table 2:** Baseline demographics, of the overall cohort and broken down into the initial cohort and secondary cohort of patients.

| Patient Demographics | Overall cohort (n=30) | Initial cohort (n=20) | Secondary cohort (n=10) |
| --- | --- | --- | --- |
| Age (years) | 68.6 ± 9.0 | 69 ± 9.0 | 67.7 ± 9.0 |
| Height (cm) | 169.1 ± 10.0 | 168.7 ± 11.0 | 169.8 ± 7.0 |
| Weight (kg) | 81.6 ± 16.2 | 80 ± 16.5 | 84.4 ± 15.6 |
| BMI (kg/m<sup>2</sup>) | 28.5 ± 4.0 | 27.8 ± 4.0 | 29.8 ± 4.0 |
| Waist-to-hip ratio | 0.96 ± 0.1 | 0.96 ± 0.1 | 0.95 ± 0.1 |
| Systolic blood pressure (mmHg) | 145.9 ± 19.6 | 147.8 ± 18.9 | 140.8 ± 22.4 |
| Diastolic blood pressure (mmHg) | 79.1 ± 8.3 | 80.6 ± 7.3 | 75.9 ± 9.4 |
| ABPI (mean of both legs) | 0.75 ± 0.2 | 0.77 ± 0.2 | 0.72 ± 0.2 |
| ICD (metres) | 118.6 ± 109.9 | 125.2 ± 119.5 | 105.4 ± 92.1 |
| MWD (metres) | 283 ± 190.5 | 293.1 ± 211.4 | 262.9 ± 148.2 |
| Gender |  |  |  |
| • Male | 23 (76.7%) | 16 (80%) | 7 (70%) |
| • Female | 7 (23.3%) | 4 (20%) | 3 (30%) |
| Smoking Status: |  |  |  |
| • Non-smoker | 2 (6.7%) | 2 (10%) | 0 (0%) |
| • Ex-smoker | 17 (56.7%) | 10 (50%) | 7 (70%) |
| • Current smoker | 11 (36.7%) | 8 (40%) | 3 (30%) |
| Respiratory disease | 6 (20%) | 4 (20%) | 2 (20%) |
| Coronary artery disease | 7 (23.3%) | 4 (20%) | 3 (30%) |
| Cerebrovascular disease | 2 (6.7%) | 1 (5%) | 1 (10%) |
| Diabetes | 8 (26.7%) | 4 (20%) | 4 (40%) |
| Statin | 28 (93.3%) | 19 (95%) | 9 (90%) |
| Antiplatelet | 28 (93.3%) | 19 (95%) | 9 (90%) |
| Optimal medical therapy | 27 (90%) | 18 (90%) | 9 (90%) |
| Antihypertensive therapy | 18 (60%) | 12 (60%) | 6 (60%) |
| Vasoactive treatment | 17 (56.7%) | 10 (50%) | 7 (70%) |
| Beta-blockade | 9 (30%) | 6 (30%) | 3 (30%) |
| BMI = Body mass index, kg/m<sup>2</sup> = kilograms per metre squared, mmHg = millimetres of mercury, ABPI = Ankle-brachial pressure index, ICD = intermittent claudication distance, MWD = maximum walking distance. Values are given as count and percentage or mean ± SD. |  |  |  |

### Feasibility, safety and tolerability

Between April 2018 and July 2019, 144 patients with IC were referred for SEP of whom 95 were eligible (66%) and 30 consented to participate in supervised HIIT (32%).

Of the initial cohort of 20 patients recruited, seven were excluded from further participation; two had abnormal ECG changes and five could not meet the criteria for a maximal effort CPET. The peak RER and percentage of age-predicted maximum HR (%) in the patient’s excluded and those meeting maximal exercise test criteria were 1.1 and 0.9 and 72% and 83% respectively (table 3). Of the remaining thirteen patients eligible and able to commence the HIIT programme, one withdrew due to an inability to tolerate the intervention, two withdrew due to developing a concurrent illness (unrelated to the study) and one withdrew due to moving out of the area. Of note, one further participant withdrew due to an adverse event probably related to the exercise intervention. This was a first isolated episode of a ‘dull chest ache’ that occurred in the period between two exercise sessions. Following referral to cardiology a diagnosis of probable angina was made but not confirmed as diagnostic angiogram was refused. Thereafter, eight of these initial twenty (40%) patients completed the supervised HIIT programme.

**Table 3:** Baseline cardiopulmonary exercise testing variables for patients in the initial cohort of 20 patients who were included or excluded on the basis of the ability to achieve maximal test criteria.

| Variable | Included (*n* =13) | Excluded (*n* = 5) |
| --- | --- | --- |
| $\dot{V}O_{2peak}$ (mL·kg<sup>-1</sup>·min<sup>-1</sup>) | 15.9 ± 3 | 12.5 ± 2 |
| Peak RER | 1.1 ± 0.1 | 0.9 ± 0.1 |
| Peak RPE | 19.8 ± 0.4 | 18.3 ± 2.1 |
| Maximum heart rate (bpm) | 126.5 ± 19.2 | 106.4 ± 13.6 |
| Percentage of age predicted maximum heart rate (%) | 82.9 ± 11.3 | 71.8 ± 10.1 |
| Peak power output (W) | 95.5 ± 33.7 | 57.3 ± 30.5 |

Values are given as mean ± SD. $\dot{V}O_{2peak}$ = peak oxygen uptake, Peak RER = respiratory exchange ratio, Peak RPE = rating of perceived exertion, bpm = beats per minute, W = Watts. N.B. Two patients excluded on the basis of ischaemic ECG changes were not included.

Following a review of the inclusion/exclusion criteria and to improve pragmatism it was decided that patients would no longer be excluded on the basis of an inability to achieve standard maximal effort CPET, but instead included and provided with the same personalised (i.e. based on their baseline CPET), time-efficient interval training programme, designed to maximise benefit. Once this exercise testing entry criterion was removed, a further ten patients were recruited of which seven (70%) completed the programme. One patient was excluded due to ischaemic ECG changes, and another withdrew three weeks after commencing the programme due to reported time constraints. One further withdrawal was due to a serious adverse event that was possibly related to the intervention. This event was a thrombosed popliteal aneurysm that occurred in the period between two exercise sessions. The patient was admitted for surgery and underwent a lower limb bypass procedure, and as such was withdrawn. Whilst it is possible that this was related to the intervention, the first symptomatic manifestation (acute limb ischemia) occurred two days after the last exercise session, meaning that it was not definitively attributable to the exercise intervention.

Of this second cohort of ten patients, two (20%) were unable to achieve a maximal effort CPET, which is comparable to the 25% in the initial cohort. Of these two patients, one completed the programme whilst the other withdrew due to time constraints. The full study processes are outlined in Figure 1.

**Figure 1.**
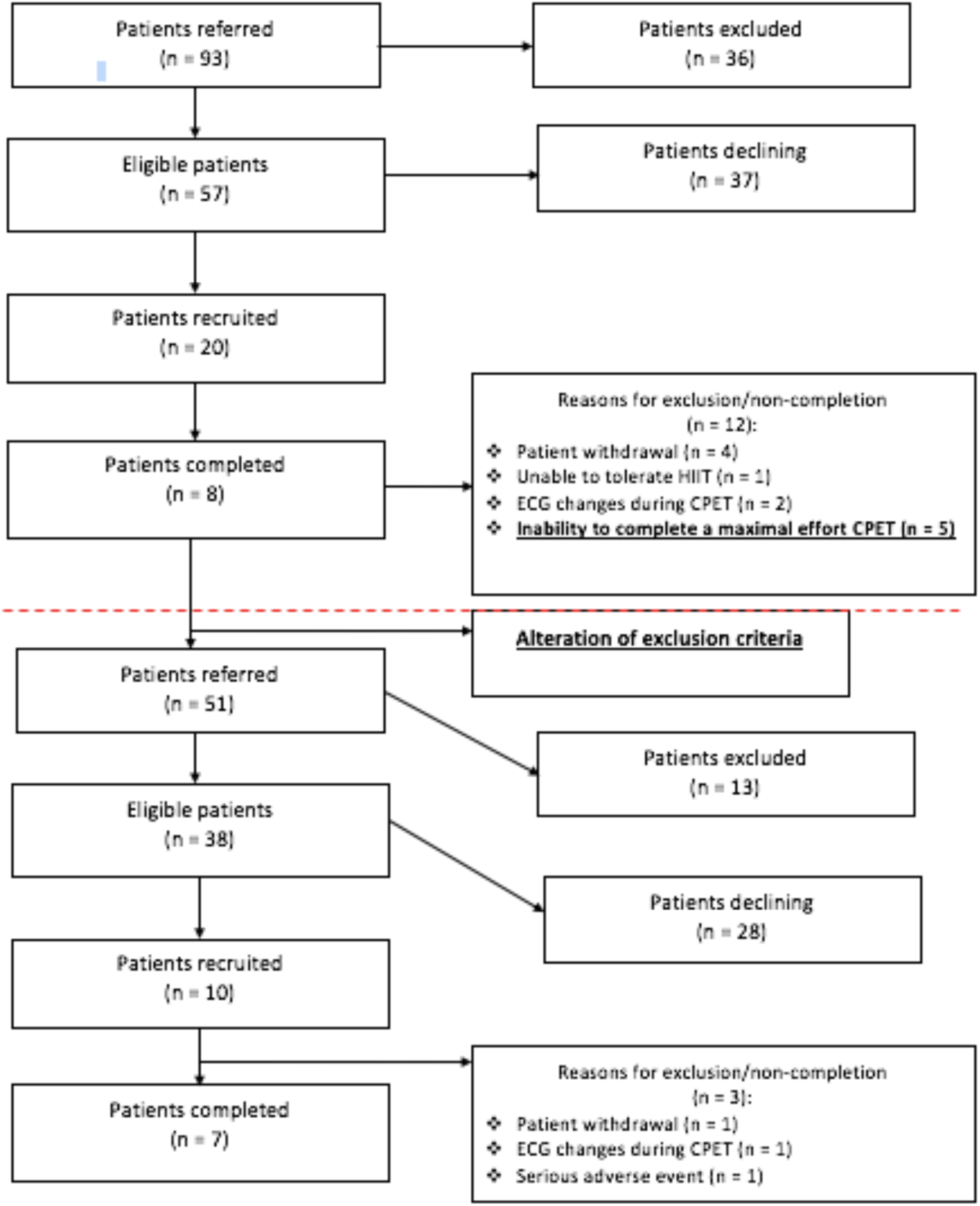
Patient flow chart.

Overall, fifteen (50%) of the thirty patients recruited in the total cohort completed the programme. Completers attended eighteen sessions, for a 100% adherence rate, over an average of 6.5 ± 0.8 weeks. ≥85%HRPeak was achieved by the second interval in 79% of sessions. All ten intervals were completed in 87% of sessions, with 14 of the 15 patients able to complete ten intervals by week two. Just one patient was unable to complete all ten intervals by the end of the programme. Two patients were withdrawn due to adverse events though neither occurred during or immediately following any HIIT exercise sessions or study visits.

### Clinical outcomes

Table 4 outlines the pre and post values for ABPI, ICD, MWD, and cardiorespiratory fitness measures for those that completed the intervention. There were improvements in ICD, MWD and peak power output.

**Table 4:** Pre- and post-intervention clinical outcomes, with mean difference for those that completed the programme.

|  |  |  |  |
| --- | --- | --- | --- |
| Variable | Pre (*n*=15) | Post (*n*=15) | Mean difference |
| ICD (m) | 114.1 ± 75.4 | 141.3 ± 62.4 | 27.3 ± 52.0 |
| MWD (m) | 243.8 ± 102.2 | 365.2 ± 153.6 | 121.5 ± 120.5 |
| Resting ABPI | 0.76 ± 0.2 | 0.78 ± 0.2 | 0.02 ± 0.1 |
| $\dot{V}O_{2peak}$ (mL·kg<sup>-1</sup>·min<sup>-1</sup>) | 15.4 ± 2.8 | 15.9 ± 3.0 | 0.46 ± 2.1 |
| VAT (mL·kg<sup>-1</sup>·min<sup>-1</sup>) | 9.8 ± 1.7 | 10.2 ± 1.7 | 0.38 ± 0.8 |
| Peak power output (W) | 94.9 ± 26.1 | 103.7 ± 30.4 | 8.8 ± 9.4 |
| Values are given as mean ± SD. ABPI = Ankle-brachial pressure index, ICD = intermittent claudication distance, MWD = maximum walking distance, VAT = Ventilatory Anaerobic Threshold, $\dot{V}O_{2peak}$ = peak oxygen consumption, Values are given as mean ± SD. W = watt, SF-36 = Short-Form 36 | | | |

### QoL outcomes

Table 5 outlines the pre and post values for QoL outcomes. There were improvements in the VascuQoL total score and a number of SF-36 domains including the physical and mental component summaries.

**Table 5:** Pre- and post-intervention QoL outcomes, with mean difference for those that completed the programme.

|  |  |  |  |
| --- | --- | --- | --- |
| Variable | Pre (*n*=15) | Post (*n*=15) | Mean difference |
| VascuQoL pain score | 4.2 ± 1.3 | 4.4 ± 1.5 | 0.2 ± 0.6 |
| VascuQoL social score | 5.5 ± 1.7 | 5.7 ± 1.0 | 0.2 ± 0.4 |
| VascuQoL activities score | 4.4 ± 1.2 | 4.6 ± 1.0 | 0.3 ± 0.6 |
| VascuQoL symptom score | 5.5 ± 0.6 | 5.8 ± 0.6 | 0.3 ± 0.6 |
| VascuQoL emotional score | 5.4 ± 1.6 | 5.4 ± 1.4 | -0.03 ± 0.8 |
| VascuQoL total score | 4.9 ± 1.1 | 5.1 ± 1 | 0.2 ± 0.5 |
| SF-36 physical functioning | 50.1 ± 18.2 | 54.7 ± 20.2 | 4.6 ± 16.3 |
| SF-36 role physical | 52.5 ± 26.9 | 55.4 ± 27.7 | 2.9 ± 18.1 |
| SF-36 pain | 47.9 ± 19.3 | 50 ± 22.6 | 2.1 ± 11.4 |
| SF-36 general health | 46.6 ± 19.9 | 52.7 ± 20.8 | 6.1 ± 15.1 |
| SF-36 vitality | 46.6 ± 19.9 | 50.9 ± 17.4 | 9.4 ± 12.8 |
| SF-36 social functioning | 76.8 ± 26.3 | 76.8 ± 27.2 | 0.0 ± 23.5 |
| SF-36 role emotional | 73.3 ± 30.2 | 80 ± 25.2 | 6.7 ± 24.2 |
| SF-36 mental health | 68.6 ± 17.1 | 73.6 ± 17.8 | 5.1 ± 14.0 |
| SF-36 physical component summary | 37.8 ± 7.6 | 39.4 ± 7.3 | 1.6 ± 5.0 |
| SF-36 mental component summary | 49.2 ± 9.9 | 51.9 ± 9.4 | 2.7 ± 6.7 |
| Values are given as mean ± SD. QoL = quality of life, VascuQoL = vascular quality of life questionnaire, SF-36 = Short-Form 36 |  |  |  |

## Discussion

The eligibility (66%) and recruitment (32%) rates for this HIIT programme were similar to those previously reported for standard SEP ^22^. With regards to completion rates, these were lower than reported within a systematic review of exercise training trials for patients with IC ^22^, but are comparable to the rates for the usual care SEP provided in our centre and likely reflective of “real-world” exercise programmes that are linked to tertiary care facilities ^44^. This suggests that HIIT may be a feasible alternative to traditional SEP for claudication and implementation of it was relatively straightforward. The patient withdrawal rate (20%) in this study is somewhat higher than that seen in HIIT programmes in other clinical populations, which are closer to 10% ^45,46^. This may be due to the small sample in this cohort, the close attention given to recording patient referrals, recruitment/withdrawals which may not be comparable within other reports and some older age and comorbidity profiles characteristics of PAD cohorts. Further, the present withdrawal rate is similar to SEPs ^22^, suggesting that HIIT is no less tolerable than current practice. In addition, the majority of sessions were completed in full and at the required intensity, further suggesting that HIIT is tolerable in this referred patient group.

There was one adverse and one serious adverse event possibly related to the HIIT intervention, though these did not require any changes to the intervention or study procedures. Notably, there were no adverse or serious adverse events that occurred during or immediately following any HIIT exercise sessions or study visits. Accordingly, this study provides preliminary findings to support the overall safety of HIIT in clinically referred IC patients, though further evidence is required. However, these early safety findings may be confounded by the use of CPET at baseline in the current study, which may help to screen out those at higher risk of a cardiac event during high intensity exercise. Current evidence suggests that CPET prior to SEP is not necessary ^47^, as the number of patients screened out is low (3.5%). Furthermore, the adverse event rate is approximated to be one event in every 10,340 patient-hours ^47^. It would be reasonable to assume that the relative intensity of SEP is lower than that for HIIT and therefore may be less likely to elicit an adverse cardiac response. Furthermore, a large proportion of PAD patients have co-existing coronary artery disease ^48^, and a number of these will be undiagnosed. Indeed, 10% of patients were excluded following a positive baseline CPET in this study. One later had confirmed triple vessel disease and underwent quadruple coronary artery bypass grafting, whilst another excluded participant required permanent pacemaker insertion. The potential cardiac risks presented by HIIT in IC patients remains largely undefined and further evidence is needed from a larger cohort of patients. Certainly, any exercise programme adopting HIIT should undertake CPET (with exercise ECG screening) prior to prescribing and performing exercise to ensure accurate exercise prescription and patient safety.

One salient finding of this study is that several patients were excluded following their baseline assessment because they were unable to achieve a maximal effort CPET. This meant that a *traditional* HIIT programme could not be prescribed. This inability to achieve a maximal test is likely to be due to severe deconditioning of patients with IC due a cycle of pain and physical activity avoidance ^6^. The mean baseline 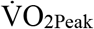 for the patients in this study was higher than previously reported in those with IC (although this may be related to the use of cycle ergometer rather than treadmill testing) at approximately 15 mL·kg^-1^·min^-1 44,49^. In comparison, a recent study considering HIIT in those with coronary artery disease reported a mean baseline value of approximately 23 mL·kg^-1^·min^-1^, whilst another study in more than 250 heart failure patients (with an ejection fraction of ≤35%) reported a baseline value of 17.1 mL·kg^-1^·min^-1 45,46^. This suggests that IC patients terminate exercise prematurely due to a perception of fatigue and/or are markedly more deconditioned than those with chronic and advanced heart disease. This may explain why a number of patients were unable to achieve a maximal effort CPET, despite subjectively feeling they have performed to their limit. Indeed, the patients unable to achieve a maximal effort test in this cohort reported a mean RPE score of >18/20.

However, those who are most deconditioned, and unable to perform a maximal effort test, have the potential to accrue the most benefit from participating in supervised, structured exercise following a personalised HIIT exercise prescription. Indeed, the VAT, a submaximal marker of cardiorespiratory fitness has been demonstrated to be a significant predictor of improvement in walking distance following SEP, with those with a lower threshold at baseline (i.e. the least fit), having the greatest improvement in walking distance ^44^.

Based on the proportion of patients who were excluded due to an inability to achieve a maximal CPET, we altered the inclusion / exclusion criteria and conducted further feasibility work. This ‘submaximal’ version of the short-term HIIT programme did not exclude patients on the basis of an inability to achieve a maximal CPET, rather we prescribed the same personalised, time efficient programme on the basis of their baseline submaximal peak workload attained. The rationale for this alteration was two-fold. Firstly, the patients excluded on this criterion, may have the potential to achieve the largest clinical benefits. Secondly, despite a similar withdrawal rate, the completion rate for the first 20 patients was just 40%. Once the maximal exercise test criterion was removed in the secondary cohort, the completion rate improved to 70%.

### Clinical and QoL outcomes

Despite not being adequately powered, the results from this study also suggest that, for patients who complete HIIT, it may provide clinical benefit in terms of walking distance and QoL. Our small sample and study design, however, precludes statistical comparison and substantive evidence of clinical benefits. Despite this, the results show promise given the improvements in MWD in this sample represent a large minimally clinically important change, whilst more importantly providing this benefit in half the usual programme duration required for standard SEPs ^50^.

As such, HIIT programmes for IC patients appear to have the potential to provide clinical and symptomatic improvements. As the intervention period is reduced from twelve to six weeks, with potentially comparable outcomes, it can reduce patient attendance burden and may be easier to deliver. This may therefore provide a cost reduction at both patient and service delivery level. Finally, this exercise prescription moves away from a *one size fits all* approach and instead adopts a personalised exercise prescription based on a CPET, with the ability to maximise patient benefit. This CPET-based exercise prescription has also recently been recommended for cardiovascular rehabilitation programmes, both in the UK and internationally ^51,52^.

## Limitations

One key limitation of this study is that participants were recruited from patients referred to a usual care SEP. It is therefore not possible to identify if patients who chose to take part in this study are simply those who would have also chosen to take part in SEP.

In addition, the small sample size precluding statistical analysis, the single-centre design and the lack of a comparison group are limitations of this study. However, this feasibility work is vital to ensure the intervention and inclusion criteria are appropriate, or whether they need to be altered, as in this case, to inform the design of future studies to assess the efficacy of this exercise intervention.

## Conclusion

This study has provided novel, but preliminary findings to suggest that patients with IC can perform a HIIT programme, with uptake and completion rates similar to standard SEPs. It has also shown that with relevant pre-screening HIIT appears safe, well tolerated and outcomes can be measured which seem to respond to the intervention. Following a small change in the exclusion criteria, it appears that the intervention and inclusion / exclusion criteria are now appropriate for this population. A further, larger proof-of-concept study appears warranted before moving to randomised controlled trials of HIIT versus usual SEP in patients with IC.

## Data Availability

Data is for descriptive, rather than statistical analysis and as such will not be provided.

## Author contributions

